# Location-allocation modeling identifies strategic health facilities to expand access to snakebite antivenom in the Brazilian Amazon

**DOI:** 10.64898/2026.08.28.26360696

**Authors:** Marcos Adriano Garcia Campos, Thiago Augusto Hernandes Rocha, Joao Vitor Perez de Souza, Letícia Murase, Felipe Murta, Marco Sartim, Jacqueline Sachett, Altair Seabra de Farias, Vinícius Machado, Fan Hui Wen, Catherine Staton, Wuelton Monteiro, Charles Gerardo, Joao Ricardo Nickenig Vissoci

## Abstract

**Background:** Snakebite envenoming is a major cause of preventable death and disability in the Brazilian Amazon, where long distances, sparse roads, and dependence on river transport delay access to antivenom. We developed location-allocation models to identify community health centers that could strategically expand access to antivenom in Amazonas State, Brazil.

**Methodology/Principal Findings:** We conducted an ecological geospatial study using a 2025 WorldPop population surface, locations of existing and candidate health facilities, and a multimodal road-and-river transportation network derived from OpenStreetMap and HydroSHEDS. Population demand was represented by 7,065 populated centroids, including 1,586 within Indigenous territories. We applied a maximize-coverage algorithm with a six-hour travel-time threshold. Two models were developed: one for Amazonas excluding Manaus and one for populations living in Indigenous territories. Both models began with 77 facilities already providing antivenom and progressively added candidate community health centers until coverage gains plateaued. The plateau occurred at 110 facilities, corresponding to 33 additional centers. In the model excluding Manaus, this configuration covered 1,118,831 people, or 75.11% of the target population; 87.61% of those covered could reach care within three hours. In Indigenous territories, coverage increased from 50.55% to 69.50%, reaching 50,434 people, of whom 81.39% were within three hours of care. Validation used 3,595 snakebite notifications from the 30 highest-burden municipalities in the Brazilian Notifiable Diseases Information System during 2023–2025. The median proportion reaching care within six hours was 40.81% in observed data and 72.17% in model estimates.

**Conclusions/Significance:** Strategically equipping 33 additional existing community health centers could substantially expand timely access to antivenom, particularly in rural and Indigenous areas. Location-allocation modeling that incorporates river transportation can support evidence-based decentralization of time-sensitive health services in geographically complex settings.

**Author Summary:** Snakebite is a serious but preventable health problem in the Brazilian Amazon. Antivenom can save lives, but many people live far from the hospitals that currently provide it, and travel often depends on rivers rather than roads. We investigated whether existing community health centers could be selected strategically to bring treatment closer to rural and Indigenous communities in Amazonas state.

We combined population maps, locations of health facilities, and road and river routes to estimate how long people would need to travel to receive antivenom. We found that adding antivenom services to 33 carefully selected community health centers, in addition to the 77 facilities already providing treatment, produced most of the achievable improvement in coverage. This arrangement could provide access within six hours for about three-quarters of the population, and nearly seven in ten people living in Indigenous territories. When we compared the estimates with snakebite notifications from 2023 to 2025, the proposed distribution showed a larger proportion of people reaching care within six hours. Our findings show how existing health infrastructure and river transportation data can guide practical decisions about where to decentralize antivenom treatment.

## Introduction

Annually more than 5 million people are bitten by snakes worldwide, with roughly 3 million cases of snakebite envenoming (SBE) and more than 100,000 deaths each year (1–3). Low- and Middle-Income Countries (LMICs) concentrate 95% of SBE cases (4). In Brazil, there are approximately 30,000 cases a year, and the Amazon region has a SBE rate 10 times greater than the Brazilian average and double that of other Amazonian regions (5).

The Brazilian Amazon region remains a neglected area, representing the highest incidence of cases per capita in the country (5). The region encompasses a diverse array of venomous snakes, in addition to reports of accidents caused by non-venomous snakes (6,7). Use of low-effective traditional healing practices, combined with the vast geographical barriers and logistical challenges of the Amazon region, continue to hinder timely and effective medical intervention (8). These factors further aggravate the situation, reinforcing the urgent need for strategies and guidelines that ensure effective access to snake antivenom for affected populations (9). Living in a more than 300□ km distance from health centers of the Amazon region is independently associated with case fatality (10).

Although antivenom treatment is public and very effective and safe in preventing deaths and complications in Brazil (11), a half of SBE patients do not receive any care from the Brazilian health system in some parts of the Amazon region (12). This implies significant socioeconomic consequences, leading to temporary or permanent work disability, lost productivity and potential healthcare costs associated with hospitalization or even mortality (13).

Solutions involving organizational designs of health services and practice arrangements based on the needs of snakebite victims and the unique characteristics of the Amazon territory can help create a decentralized scenario of SBE treatment with antivenoms (14,15). Decentralization programs aimed at prevention and treatment of SBE in remote areas in the Amazon region have already proven effective (16). Innovative ways to reduce the time between the SBE victim and the care center provided with antivenom have already been explored with good results, such as the use of rivers for transportation (17).

The aim of this study is to develop a location-allocation model (LAM) to optimize antivenom distribution in the Brazilian Amazon region using decentralized health facilities as providers centers to antivenom administration.

## Methods

### Study Design

We conducted an theoretical ecological study using publicly available secondary data from the Amazonas state, the largest Brazilian state in the Amazon region. Through the geographic information system (GIS) we employed network analysis and LAM to compute realistic travel times, delineate service catchments, and evaluate facility parameters that maximize population coverage. Three domains structured the analysis: Demand (population density and distribution); Supply (location and type of health facilities); Mobility (distance between population and health facilities). The selection of the most strategic health facilities is done automatically by the LAM. All variables were calculated and mapped on the ArcGIS Pro 3.1.2. Only aggregated, non-identifiable data were used, and no individual-level inferences were made. All the analysis workflow is represented in Figure 1.

**Figure 1.**
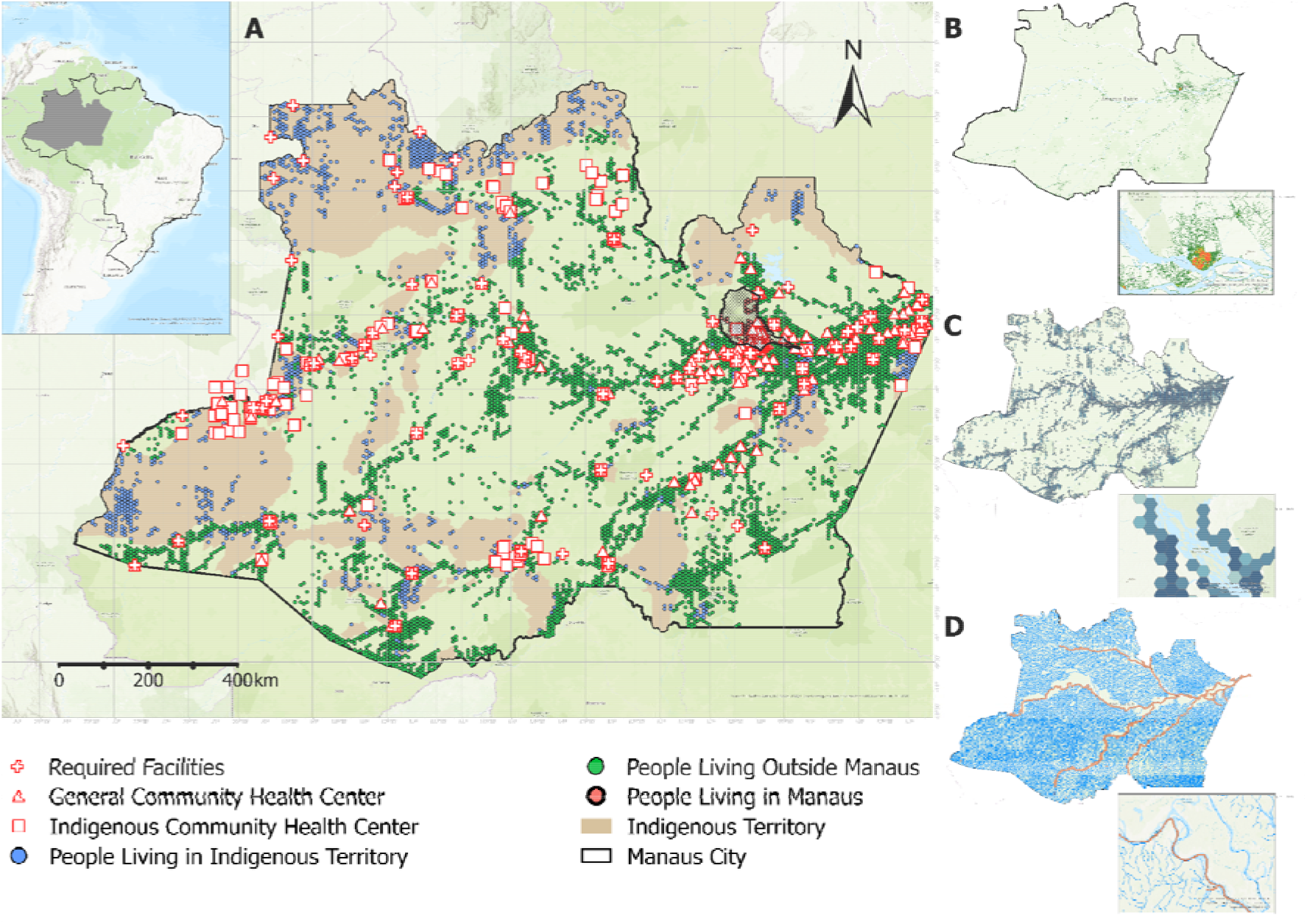
Development of the location-allocation model (LAM) scenario for identifying healthcare facilities for snakebite envenoming treatment. **A:** Map of Amazonas State, located in the Brazilian Amazon region (inset), showing the geolocation of 77 required facilities that currently provide antivenom, 633 community health centers (primary care facilities), and 217 Indigenous community health centers serving Indigenous populations. The scattered green and blue points represent the spatial distribution of the population, including people living in Indigenous territories. **B:** Population density in Amazonas state according to WorldPop, demonstrating that most of the population is concentrated along major rivers. **C:** A grid of 50-km^2^ hexagons, with each hexagon containing the total population within its area; different shades of blue represent different population densities. A centroid was generated for each hexagon and used as the population demand point connected to healthcare facilities in the LAM. **D:** Navigable rivers derived from HydroSHEDS, shown in blue, and commercial river routes obtained from OpenStreetMap. These spatial datasets and transportation networks formed the analytical framework used to develop the LAM.

### Defining Demand

The demand was represented by the dasymetric population living in Amazonas state. To estimate the number of inhabitants within the setting area, we used the raster file from WorldPop dataset corresponding to the most recent 2025 dasymetric population surface at 100-meter resolution, which calculates the sum of the pixel values corresponding to the population per square kilometer (18). For each analysis unit, totals were obtained by summing the pixel values (people per pixel) (19). WorldPop uses a top-down dasymetric approach to redistribute official census counts through machine-learning models based on remotely sensed and ancillary covariates, preserving administrative totals while generating high-resolution, spatially explicit population estimates (20).

The current Brazilian census does not show the spatial distribution of the population. It was generated in 2022 by the Brazilian Institute of Geography and Statistics (IBGE, abbreviated in Portuguese) and estimated the population of the state of Amazonas at 3,941,613 people (21). WordPop estimated a population for the same region at 3,134,646 people. Shapefile maps of the studied territory were extracted from the official IBGE website and used to delimit the area of interest in the GIS software (22).

After obtaining a map showing the spatial distribution of the population of the state of Amazonas, a 50 m^2^ hexagonal grid layer was overlaid, and the population contained within each polygon was aggregated. The hexagonal shape has been used to some advantage, primarily to better fit the irregular boundaries of territories (23), such as the state of Amazonas. The total population clipped within each hexagon was then assigned to the centroid of that hexagon. We excluded all points representing hexagonal areas with no population. In the proposed models, each point represented a reference spot to be connected to the health facilities. In total, 7,065 points were generated across the state of Amazonas, of which 1,586 were located within Indigenous territories.

### Supply Features

Two types of supply facilities were considered: health services that currently administer antivenom provided by the Butantan Institute, the main distributor of antivenom in Brazil, in the public health system of the state of Amazonas (called required facilities); and services with the potential structure to administer antivenom (called candidate facilities). We found 77 services provided with antivenom according to Butantan Institute (24).

For the candidate facilities, we considered the primary care health units in the state of Amazonas. Despite having less robust infrastructure than tertiary hospitals, they have already proven to be safe for administering antivenom in remote regions of Amazonas (16). In total, there are 633 Community Health Centers (CHCs) scattered throughout the municipalities of Amazonas state, offering public health services in a decentralized system. For the Indigenous population, 217 health facilities specializing in their care (Indigenous-CHC) were also included in the model as candidate facilities.

### Calculating Mobility

We modeled mobility using two open-source line shapefiles: OpenStreetMap (OSM) and HydroSHEDS. From OSM, we extracted streets, highways, and fluvial waterways within the state of Amazonas. OSM is a community-curated, open geospatial database where volunteers and organizations map the world’s transportation and settlement features (25). Its data are continuously updated and can be freely used and edited under an open license. This makes OSM a suitable, transparent source for detailed road geometry and attributes across urban and rural areas (26).

For river travel (widely used in Amazon Region), we used the HydroSHEDS river network to represent navigable rivers in Amazonas. HydroSHEDS (“Hydrological data and maps based on SHuttle Elevation Derivatives at multiple Scales”) is a global hydrographic dataset derived primarily from satellite elevation data (27). It provides hierarchically organized river networks and drainage information, enabling hydrologically consistent extraction of channels and identification of main stems versus tributaries, capabilities that are essential in river-based accessibility modeling in the Amazon (28). Information on navigable rivers was based on a previously published and validated study for the same analysis region (17).

Using these two line layers, we built a multimodal (road and river) network that connects all routes topologically (29). For each edge, we computed a travel-time attribute to enable cost-based routing. Travel time in hours was calculated as the segment length (in kilometers) divided by the maximum travel speed for that segment (in kilometers per hour). Segment length and maximum travel speed are variables available in the original databases.

### Adjusting Parameters

We used the LAM analysis based on the network transportation file created. The LAM is a spatial optimization technique designed to identify the most suitable locations for facilities in relation to a given demand. It considers the geographic distribution of the population, the transportation network, and accessibility to optimize coverage and minimize travel distance (30).

The Maximize Coverage algorithm was applied to select facilities in a way that maximizes the number of people served within a specified travel distance or time. This approach prioritizes the attractiveness of facilities closer to the demand, simulating real-world behavior in which individuals typically seek the most accessible service option (31).

We defined the demand toward the facility time in a cut-off of six hours (28). Administered antivenom within six hours is considered the golden window-time to prevent and reverse envenoming complications of the SBE (33,34).

The number of chosen facilities was gradually studied based on the population coverage. Starting with 78 facilities (all required facilities and at least one candidate), we added one by one facility and described the number of people covered by the model. The optimal number of chosen facilities was defined as a plateau point where additional facilities yield no significant increase in population coverage. At the end, the model was able to show the strategic location of the chosen facilities and the best routes between demand and the facility points.

### Designing Models

Two distinct models were proposed. The first considered the entire population of the state of Amazonas, excluding the population of the capital, Manaus, the state’s most populous city. Manaus was excluded because it is already fully covered within the defined travel-time threshold; therefore, the model was designed to assess less populous and more remote or rural areas. The second model was specifically tailored to Indigenous territories in the state of Amazonas. For this population, all CHC and Indigenous-CHC were considered candidate health facilities.

The Indigenous population was defined as individuals living within the Indigenous territories of Amazonas. Shapefiles for these territories were obtained from online data provided by the National Foundation for Indigenous Peoples (FUNAI, in the Portuguese acronym) (35).

### Validation

To validate the proposed LAMs, we compared municipality-level travel-time distributions estimated by the models with observed time to care among snakebite patients recorded in the Brazilian Notifiable Diseases Information System (SINAN, in Portuguese abbreviation) from 2023 to 2025. The SINAN dataset included accidents involving both venomous and nonvenomous snakes in Amazonas state. The 30 municipalities with the highest number of snakebite notifications during 2023-2025 were selected, excluding Manaus city to focus on rural and remote areas. Corresponding LAM estimates were then extracted for these same municipalities. SINAN also includes a variable identifying Indigenous ethnicity, which was used in the validation of the Indigenous-population model (36).

For both SINAN and the LAM estimates, time to care was classified as 0–6 hours, >6–12 hours, or >12 hours. In SINAN, time to care was defined as the interval between the snakebite and attendance at a health service, whereas in the LAM it represented the estimated travel time from population points to the selected CHCs. For each municipality, we calculated the percentage of patients or modeled population within each interval. Comparisons between SINAN and LAM were paired by municipality and summarized using means, standard deviations, medians, and interquartile ranges (IQR). Differences between the two sources were assessed using the paired Wilcoxon signed-rank test, with statistical significance defined as a two-sided p-value <0.05. All statistical analyses and graphics were generated using R version 4.6.1.

## Results

The estimated population of Amazonas state was 3,134,646 inhabitants, closely aligned with census estimates. Of this total, 1,489,590 people were estimated to live in the non-capital areas, and 72,567 in Indigenous territories. The progressive addition of CHCs increased population coverage until a plateau was reached at 110 facilities in both LAMs (Figure 2). This setting comprised the 77 required CHCs plus 33 additional CHCs selected by the models. In the first model, the 110-CHC chosen covered 1,118,831 people, corresponding to 75.11% of the target population and an increase of 14.12% compared with the scenario containing only the 77 required facilities. Although the maximum travel-time threshold was 6 hours, 87.61% of the covered population could reach a CHC within 3 hours.

**Figure 2.**
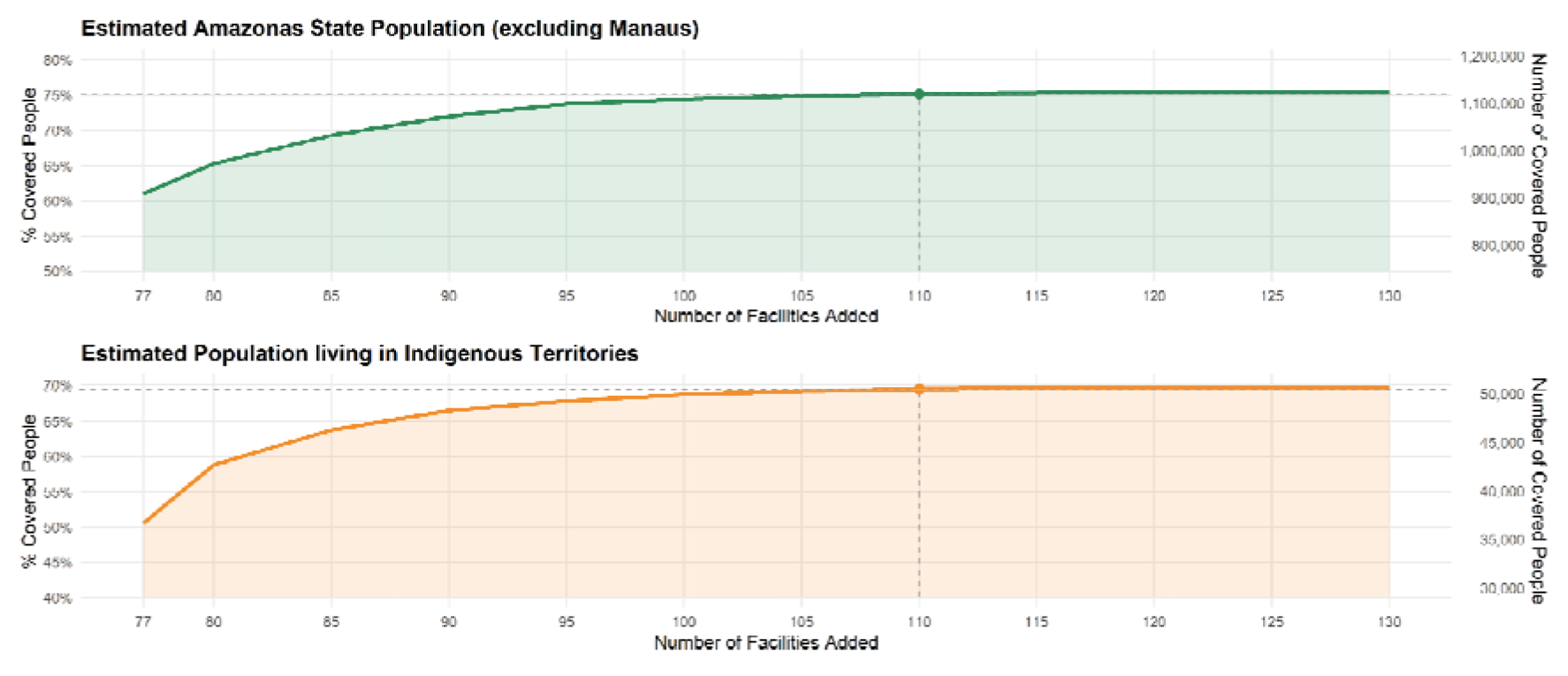
Identification of the population-coverage plateau for snakebite envenoming treatment estimated by the location-allocation model (LAM). The graphs show the absolute and relative population coverage achieved as additional healthcare facilities were incorporated into the model. Both analyses began with 77 required facilities that currently provide antivenom. Coverage increased progressively until reaching a plateau at 110 facilities, after which the addition of further services produced only minimal gains. The same plateau point was observed for the population living in Indigenous territories, shown in the lower panel.

For the population living in Indigenous territories, coverage increased from 50.55% with the 77 required CHCs to 69.50% with 110 facilities, corresponding to 50,434 covered individuals. Among the covered Indigenous population, 81.39% could reach a CHC within 3 hours. The spatial distribution of the additional CHCs identified as the most strategic by the LAMs is presented in Figure 3.

**Figure 3.**
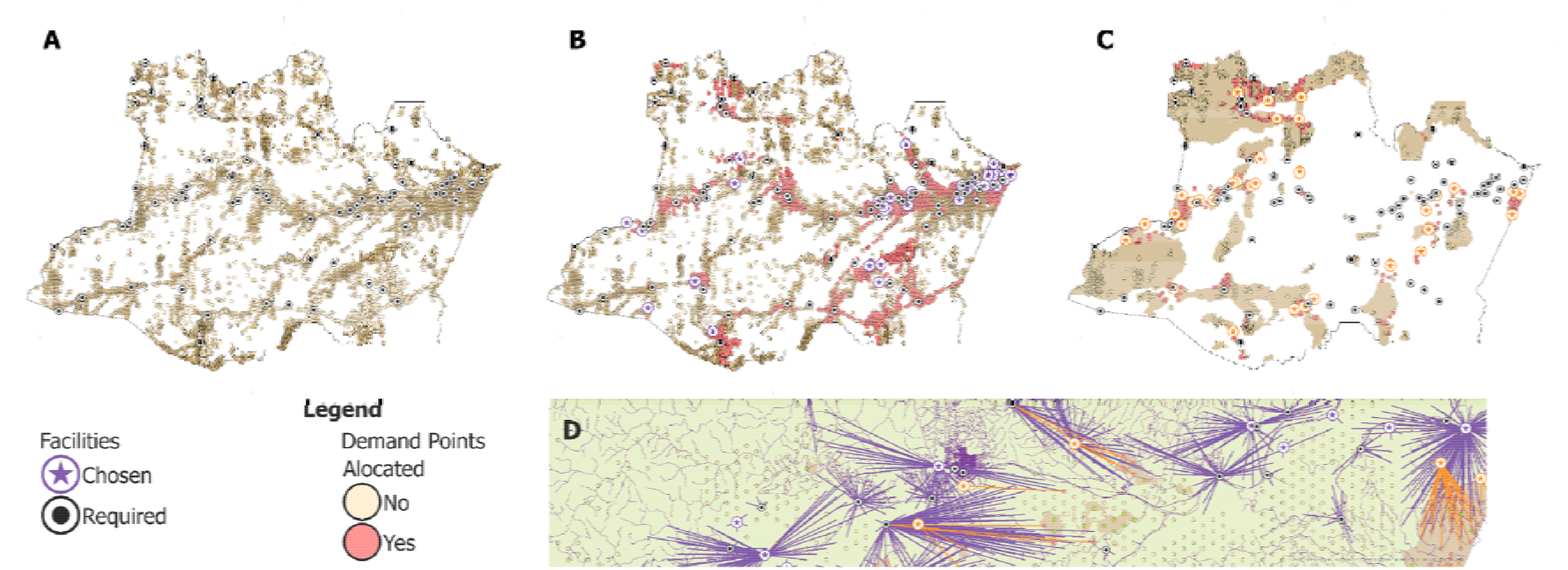
Selection of the most strategic healthcare facilities by the location-allocation model (LAM). **A:** Spatial distribution of the required facilities, showing their concentration in areas of higher population density, represented by ochre shading, while several populated areas remain uncovered. **B:** Distribution of the 110 selected healthcare facilities, comprising required facilities and community health centers (CHCs), with the allocated population shown in red. **C:** Location of CHCs serving Indigenous populations, represented by circles containing an orange star, and Indigenous territories are delimited on the map. **D:** Population demand points connected to their assigned healthcare facilities. Although these connections are displayed as straight lines, the allocation was based on travel through the actual transportation network, accounting for navigable river routes, travel speeds, and natural barriers along each route.

Between 2023 and 2025, SINAN recorded 5,587 snakebite cases involving venomous and nonvenomous snakes in Amazonas state, including 1,356 cases among Indigenous patients (24.27%). After excluding Manaus, restricting the analysis to patients who reached healthcare within 24 hours, and selecting the 30 municipalities with the highest number of notifications, the validation sample included 3,595 cases. Of these, 968 occurred among Indigenous patients (26.93%). The mean number of reported snakebite cases was 119.83 ± 60.88 cases per municipality.

Across the selected municipalities, the median proportion of patients who reached healthcare within 6 hours according to SINAN was 40.81% (IQR: 38.73–43.97%). For the same municipalities, the median proportion estimated by the LAM to reach the chosen CHCs within 6 hours was 72.17% (IQR: 47.46–83.01%), as shown in Table 1. The LAM therefore estimated a higher proportion of individuals reaching care within 6 hours. The proportions in the >6–12-hour interval were similar between the two sources, whereas SINAN showed a higher proportion of patients requiring more than 12 hours to reach healthcare. The selected municipalities and their corresponding LAM or SINAN estimates are presented in Figure 4.

**Table 1.** Validation of the model. SINAN (Brazilian Notifiable Diseases Information System) provides observed data from 2023–2025 on the time elapsed between snakebite occurrence and arrival at a healthcare facility among patients residing in the 30 municipalities of Amazonas state, Brazil, with the highest number of reported snakebite cases. The location-allocation model (LAM) estimated travel times from population points to strategically selected Community Health Centers (CHC) in these same municipalities. The table presents the municipality-level percentages of patients or modeled population within each time interval. A higher proportion reaching a CHC within 6 hours and a lower proportion requiring more than 12 hours in the LAM indicate shorter estimated access times compared with those reported in SINAN.

| **Time to reach health services** | **Model** | **Mean percentage of patients** | **Standard Deviation** | **Median percentage of patients** | **Q1** | **Q3** |
| --- | --- | --- | --- | --- | --- | --- |
| 0 to 6 hours | SINAN | 41.80 | 4.29 | 40.81 | 38.73 | 43.97 |
|  | LAM | 58.60 | 28.28 | 72.17 | 47.46 | 83.01 |
| > 6 - 12 hours | SINAN | 5.05 | 3.03 | 5.96 | 4.09 | 7.35 |
|  | LAM | 11.16 | 13.54 | 8.16 | 2.82 | 14.05 |
| > 12 hours | SINAN | 53.15 | 2.15 | 52.78 | 52.02 | 54.44 |
|  | LAM | 30.24 | 26.91 | 14.86 | 5.15 | 40.66 |

SINAN: Brazilian Notifiable Diseases Information System; LAM: Location-Allocation Model; Q1: first quartile; Q3: third quartile.

**Figure 4.**
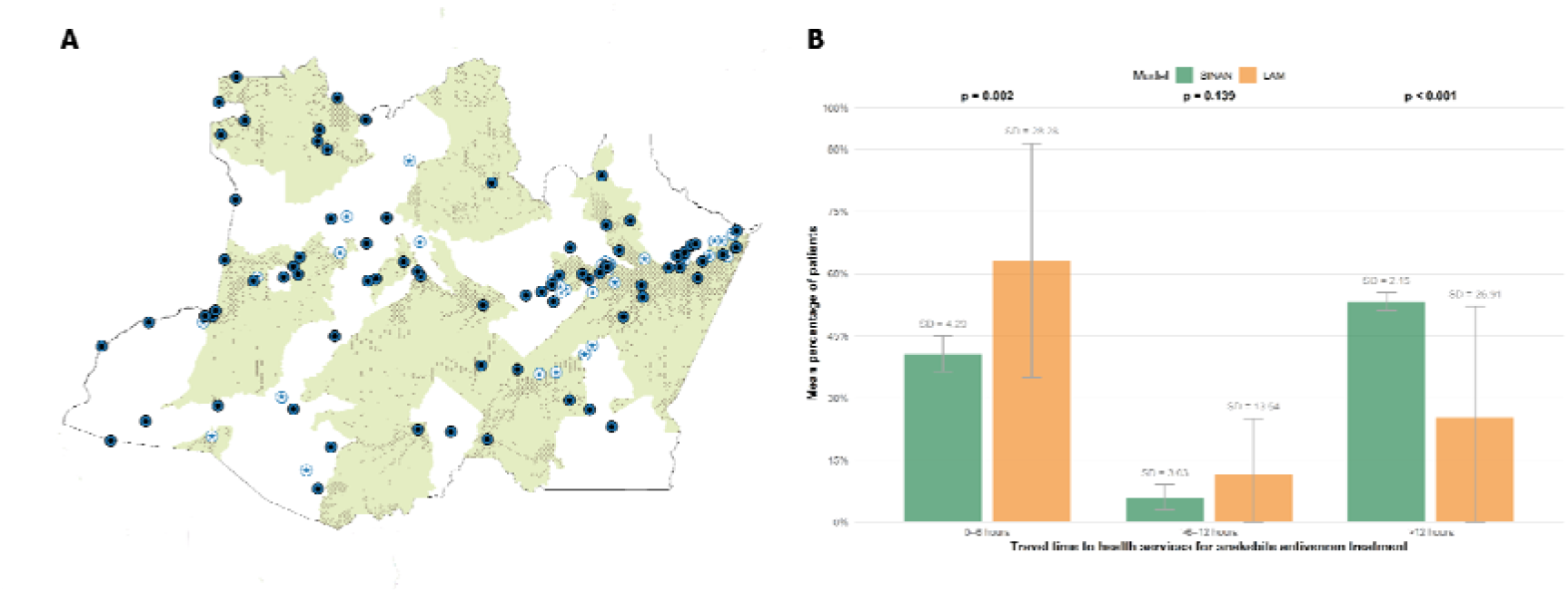
Validation of the location-allocation model (LAM) using observed data from the Brazilian Notifiable Diseases Information System (SINAN). **A:** Areas highlighted in green represent the 30 municipalities with the highest number of reported snakebite cases in Amazonas State between 2023 and 2025. Population distribution within each municipality is represented by small black points. The map also shows the 110 healthcare facilities included in the LAM: required facilities that currently provide antivenom are represented by symbols containing a black circle, whereas additional CHCs selected by the model are represented by symbols containing a blue star. **B:** Bar chart comparing the mean municipality-level proportions observed in SINAN and estimated by the LAM across different time intervals. The LAM estimated a higher proportion of individuals reaching a healthcare facility within 0–6 hours and a lower proportion requiring more than 12 hours, suggesting that the proposed distribution of facilities could improve timely access to antivenom treatment compared with historically observed access patterns.

## Discussion

This study developed LAMs to identify strategic CHCs for decentralizing antivenom treatment in Amazonas. Adding 33 CHCs to the 77 existing facilities increased six-hour coverage to 75.11% outside Manaus and to 69.50% in Indigenous territories. Compared with real-world data from SINAN, the LAM estimated a higher proportion of individuals reaching care within six hours, and fewer requiring more than 12 hours. These findings suggest that targeted decentralization could reduce geographic inequities in access to antivenom.

SBE remains a neglected tropical disease that disproportionately affects rural, Indigenous, and socioeconomically vulnerable populations (37). Despite the availability of an effective treatment, preventable deaths, disabilities, and complications continue to occur because antivenom is frequently unavailable near the communities at greatest risk (38,39). The World Health Organization emphasizes that antivenoms should be accessible through primary healthcare services in areas where snakebites occur, as delays in administration contribute substantially to unfavorable outcomes (40). In the Brazilian Amazon region, a high proportion of patients take more than three hours to reach healthcare, and prolonged access time has been associated with greater occurrence of moderate and severe SBE (37). Therefore, addressing snakebite in the state of Amazonas requires not only adequate antivenom production and procurement but also public policies capable of improving its geographic distribution (41).

In this context, LAM can provide a potentially resource-efficient decision-support tool for health planning (42). Rather than proposing the construction of an entirely new network of facilities, our approach identifies existing CHCs that could generate the greatest improvements in population coverage if equipped to provide antivenom. The use of open health, population, hydrographic, and road-network data also increases the reproducibility and potential scalability of the method. Consequently, the model may help policymakers prioritize investments in facilities where training, antivenom stocks, refrigeration equipment, communication systems, and referral capacity are likely to produce the greatest population benefit (43). Nevertheless, although this approach may optimize the use of existing resources, a formal economic evaluation is needed before it can be described conclusively as cost-effective.

The proposed decentralization is consistent with the recognition that antivenom treatment does not necessarily need to remain restricted to high-complexity hospitals. Recent initiatives in the Brazilian Amazon have demonstrated the feasibility of administering antivenom in selected Indigenous-CHC when minimum requirements for infrastructure, trained personnel, medication storage, emergency management, and referral are met (16,44). The SAVING Program, for example, has provided an important implementation framework for decentralizing treatment to Indigenous health units in Amazonas (16). Its experience also demonstrates that decentralization must extend beyond the physical provision of antivenom and include professional training, maintenance of the cold chain, availability of essential medicines and equipment, clinical protocols, continuous supervision, case notification, and well-defined referral pathways (16). Thus, the CHCs identified by our model should be interpreted as priority locations for further assessment rather than facilities that are immediately prepared to administer antivenom.

A major contribution of this study was the incorporation of river networks into the accessibility analysis. In the Amazon region, rivers are not merely geographic features but form the principal transportation network for many remote and riverine communities (45). Previous research involving Indigenous communities in the western Brazilian Amazon has identified the lack of motorized transport, financial resources, and navigable routes as important barriers to receiving hospital treatment following snakebite (46). By combining roads and hydrography, our multimodal network provides a more realistic representation of regional mobility than analyses based exclusively on terrestrial routes or straight-line distance. This approach is particularly relevant in a state where road networks are sparse and access to municipal centers frequently depends on river travel.

Nevertheless, even after optimization, universal coverage within six hours was not achieved. This result reflects the vast territory of Amazonas, its low population density, the dispersed distribution of settlements, and the extreme remoteness of several Indigenous and riverine communities. In some locations, the distance between communities and existing health infrastructure may remain too great for decentralization to fixed CHCs alone to ensure timely access (47). River transportation is also affected by seasonal water-level variation, navigability, weather conditions, boat availability, fuel costs, engine power, and waiting time before departure (48). These factors were not fully represented in the model and may produce substantial differences between theoretical and actual travel times. Consequently, complementary strategies may be required in areas that remain uncovered, including emergency river transport, mobile health units, strategically positioned transport bases, communication systems, and community-based first-response protocols.

The comparison with SINAN data provides supportive evidence for the potential benefit of the proposed allocation. However, these measures represent related but not identical concepts. The model estimates travel through the available transportation network, whereas the SINAN interval between the bite and hospital admission may also include the time required to recognize the severity of the SBE, seek assistance, arrange transportation, undergo intermediate transfers, or initially use traditional treatments. Therefore, the shorter times generated by the models should not be interpreted as a direct prediction of the time that would be observed after implementation. Rather, they represent the reduction in geographic travel burden that could be achieved if the selected facilities became operational and patients were able to access them without substantial additional delays.

Other limitations should also be considered. Population demand was estimated from a modeled population surface rather than individual residential locations, and the accuracy of the results depends on the completeness of the population, facility, road, and hydrographic datasets. Travel speeds and network connections may not capture local transportation conditions or seasonal changes in river routes. In addition, the six-hour threshold simplifies a clinical process in which earlier antivenom administration is generally preferable. The model also assesses geographic accessibility but does not directly incorporate facility readiness, workforce availability, local snakebite incidence, antivenom consumption, cultural acceptability, or the capacity to manage adverse reactions and severe complications. These factors should be evaluated before selecting facilities for real-world implementation.

This study provides a robust and reproducible framework for improving access to antivenom treatment in geographically complex and resource-constrained settings. By integrating LAM, open datasets, GeoAI-derived population surfaces, and multimodal transportation networks that recognize rivers as essential routes in Amazonas, the proposed approach identifies strategic opportunities to decentralize antivenom delivery through existing CHCs. Although important geographic gaps would remain, particularly among Indigenous and remote populations, the findings demonstrate that a targeted reorganization of current health infrastructure could substantially expand timely access to treatment. Beyond supporting antivenom decentralization policies, this methodology offers a valuable basis for future implementation, and may also guide the allocation of other time-sensitive health services where detailed population and mobility data are limited.

## Data Availability

All data produced in the present study are available upon reasonable request to the authors.

## Acknowledgments

This study was supported by the Josiah Charles Trent Memorial Foundation Endowment Fund and the National Council for Scientific and Technological Development (CNPq).

